# The Histological Diagnosis of Breast Cancer by Employing scale invariant ResNet 18 With Spatial Supervised Technique

**DOI:** 10.1101/2021.09.06.21263185

**Authors:** Syed Usama Khalid Bukhari, Asmara Syed, Syed Safwan Khalid, Syed Sajid Hussain Shah

## Abstract

**Background:** Breast cancer is one of the most prevalent cause of morbidity and mortality in women all over the world. Histopathological diagnosis is a vital component in the management of breast cancer. The application of artificial intelligence is yielding promising results for the better patient care.

**Aim:** The main aim of the present research project is to explore the potential of spatial supervised technique to develop scale invariant system for the histological diagnosis of breast cancer.

**Materials and Methods:** The anonymized images of hematoxylin and eosin stained section of the dataset, which has been acquired from the website. The slides were taken at different zoom (magnification) levels. Spatial supervised learning has been employed to make a scale invariant system. We used 400x and 40x to generate the results. For the 400x, we trained our network on a dataset of 200x,100x, and 40x images. The datasets were split into training and validation sets. The training set contained 80% digital slides of the respected dataset, and the validation set contained 20% digital slides of the respected dataset. The final result was generated by splitting the dataset of 400x into the training and test dataset. The training set contained 50% digital slides, and the test set also contained 50% digital slides. This unusual split is done to show how good spatial supervised learning works. Similarly, for 40x, we trained our networks on a dataset of 400x,200x, and 100x. The same steps were followed to obtain the 40x results.

**Results:** The result analysis revealed that the ResNet 18 with spatial supervised learning on dataset of 40x yielded the F-1 score of 1.0, while ResNet 18 with supervised learning only, on dataset of 40x yielded F-1 score of 0.9823. ResNet 18 with spatial supervised learning on dataset of 400x revealed F-1 score of 0.9957, and ResNet 18 with supervised learning only, on dataset of 400x showed the F-1 score of 0.9591. For supervised learning dataset is spited into training (80%) and testing (20% of dataset).

**Conclusion:** The analysis of digitized pathology images with the application of convolutional neural network Resnet -18 architecture with spatial supervised learning revealed excellent results, which is demonstrated by a very high F-1 score of 1.0.

The development of scale invariant system with application of spatial supervised technique solved the problem of images with variable magnifications. The finding would further pave the pathway for application of deep learning for the histological diagnosis of pathological lesions.

## Introduction

Breast cancer is the most common malignant tumor in women [1]. It is one of the most important causes of morbidity and mortality all over the world. Breast cancer is the most common cause of cancer deaths in female. In 2018, the estimated number of new cases of breast cancer is more than two million [2]. The estimated number of deaths due to breast cancer in 2018 is more than six hundred thousand [2].

Breast carcinoma most frequently occurs in old age. The mean age of breast cancer patients is 47 to 49 years [3,4] The risk factors of breast cancer include family history, reproductive history, advancing age and excessive intake of saturated fat [5]. The early menarche, low parity, lack of breast feeding and old age at first pregnancy also increase the risk of breast malignancy [6,7].

Painless lump is the most frequent clinical presentation of breast cancer. Mammogram is usually performed initially for the assessment of breast mass. The definitive diagnosis of breast carcinoma depends on the microscopic identifications of malignant cells which may be performed by fine needle aspiration cytology or biopsy. The classification of breast cancer is more accurately done by histological examination in which the cytological features of malignancy such as hyperchromasia and pleomorphism are noted along with the pattern of arrangement of cancer cells. Since the histological diagnosis of breast cancer is a very critical because the decision of major surgery depends on it. To provide assistance to histopathologists for the improvement in the accuracy and speed of histopathological diagnosis, the use of artificial intelligence needs to be explored.

The aim of the present research is to find out the accuracy of ResNet 18 for the diagnosis of breast carcinoma by evaluating the digital images of hematoxylin and eosin stained sections.

## Materials & Methods

After getting the approval from the institutional research ethical committee, the research project is performed on the anonymized images of hematoxylin and eosin stained section from the dataset. The dataset of histopathological digital slides has been acquired from the Kaggle website [8]. The slides were taken at different zoom (magnification) levels. These levels are 40x, 100x, 200x, and 400x, which are with the magnification of 40, 100, 200 and 400 respectively and these contained two categories labeled as benign and malignant.

A pre-trained ResNet -18 on ImageNet is used [9]. The following steps were performed to acquire the results. We froze the first eight (8) layers of ResNet-18, as they mostly contained the filters related to line and edge and curve detection. We removed the last fully connected layer and changed it with our own fully connected layer with five hundred and twelve (512) inputs and two (2) outputs. The network is trained on a 400x dataset of digital slides for ten epochs. The network’s weights were saved, and then the saved weights network is trained on 200x and 100x dataset, respectively. For the training purpose, a dataset of 400x, 200x, and 100x were split into training and validation sets. The training set contained 80% digital slides of the respected dataset, and the validation set contained 20% digital slides of the respected dataset.

After training the network on 400x,200x, and 100x we used the same weights to train the 40x dataset, but this time beside the last layer, all weights of the remaining layers were frozen. 40x dataset was split into training and test dataset. For training purposes, we allocate 50% of digital slides; similarly remaining 50% of the digital slides of the dataset were put in the testing dataset. The test dataset is always less than the training dataset in the convention, but here we deliberately make them equal to prove that our method (spatial supervised technique) gives good results even if the train is on a smaller dataset.

A standalone program is developed, where a pre-trained ResNet-18 on ImageNet, is used to train the 40x dataset. This time dataset is split into an 80:20 ratio; 80% of the dataset for training purposes and 20% for testing purposes.

A comparison of results by both techniques is presented in the result section. An inverse of the above mention method is also done and presented in which datasets of 40x,100x, and 200x were used to train using Spatial Supervised Technique (SST), and 400x dataset is used as final prediction set. The result of this inverse is also given in the result section.

## Results

In the present study, the digitized pathology images have been analyzed by the convolutional neural network Resnet -18 architecture with spatial supervised technique. ResNet 18 with SST on 40x revealed excellent results without any error in the correct diagnosis of the digital images of both lesions. The F-1 score with this technique is 1.0. The result is depicted in figure 1. The second technique used in this study is ResNet-18 with supervised learning on 40x, which revealed the F-1 score of 0.9823. The results are shown in table 2 and figure 2. The third category in this study is to evaluate the ResNet-18 with SST on 400x (magnification of 400). In this category, the test results revealed F-1 score of 0.9957. The results are present in table 3 and figure 3. The fourth category is ResNet 18 with supervised learning on 400x which revealed the F-1 score of 0.9591. The results are summarized in figure 4 and table 4.

**Fig1.**
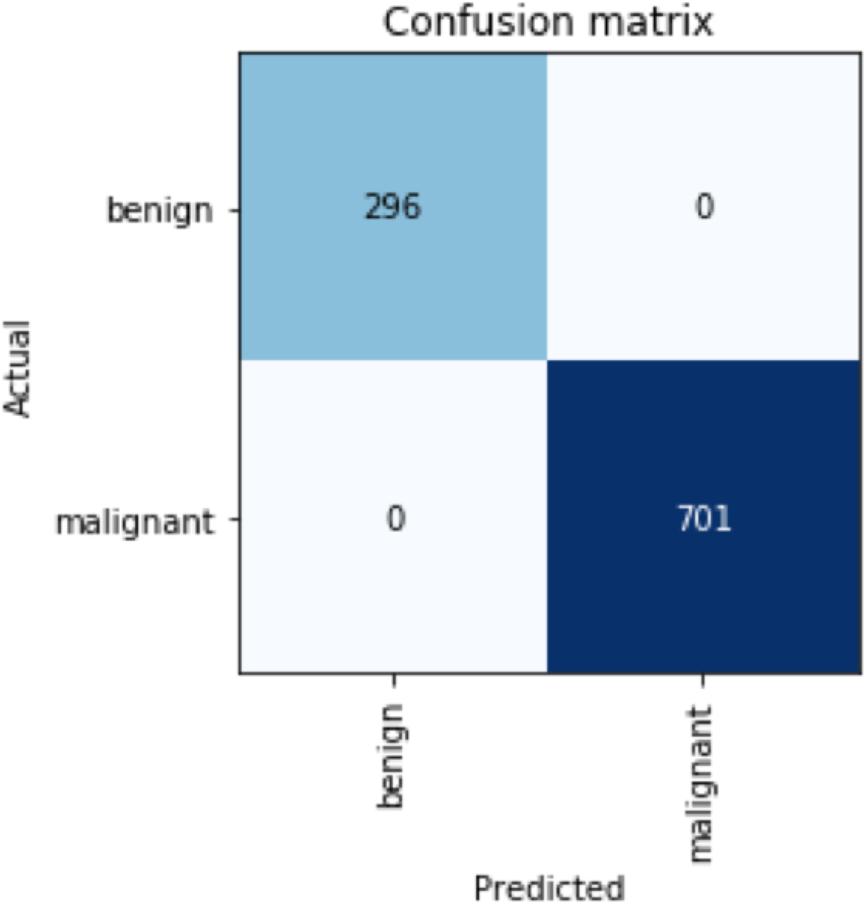
ResNet 18 with SST on 40x

**Table 1.**
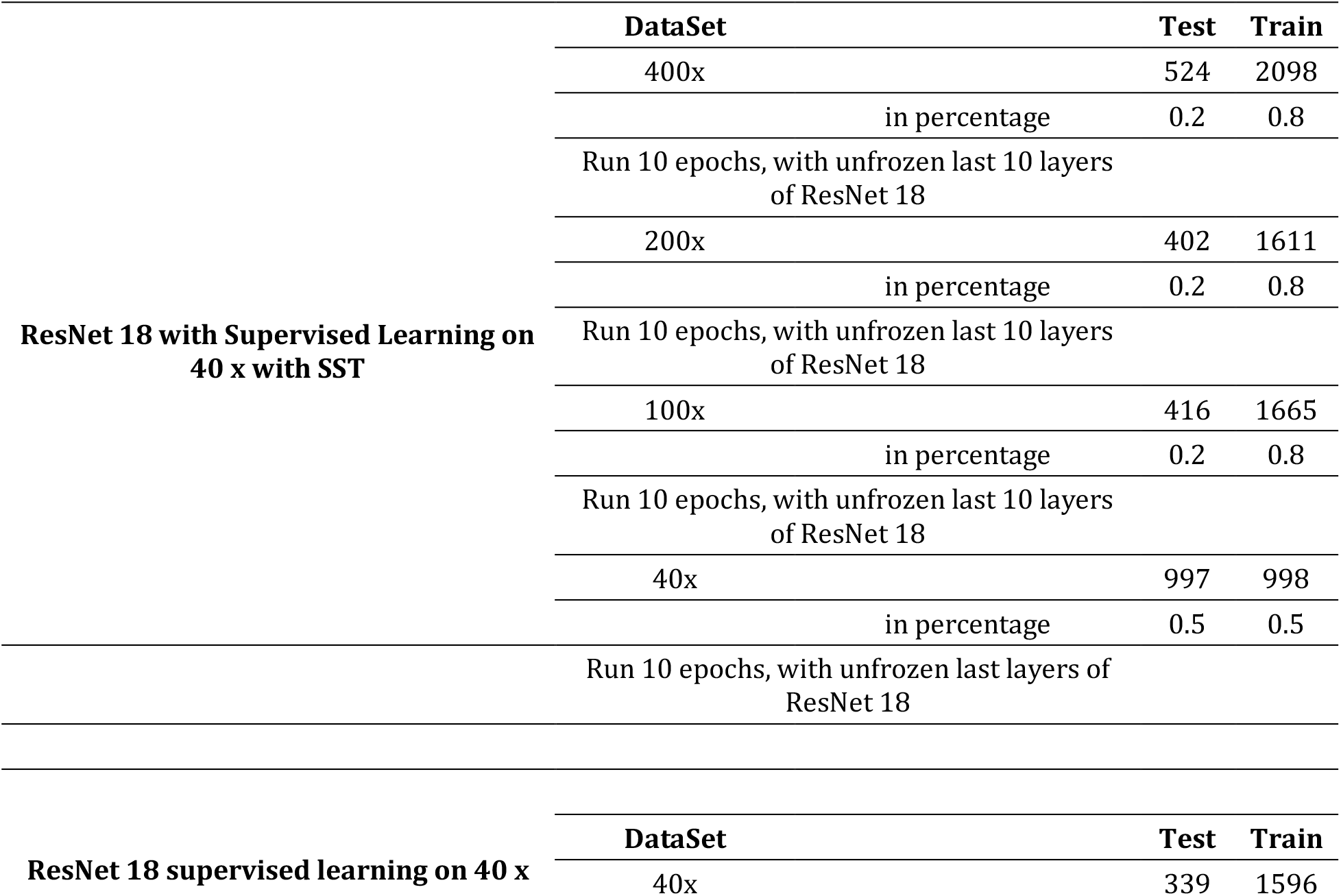
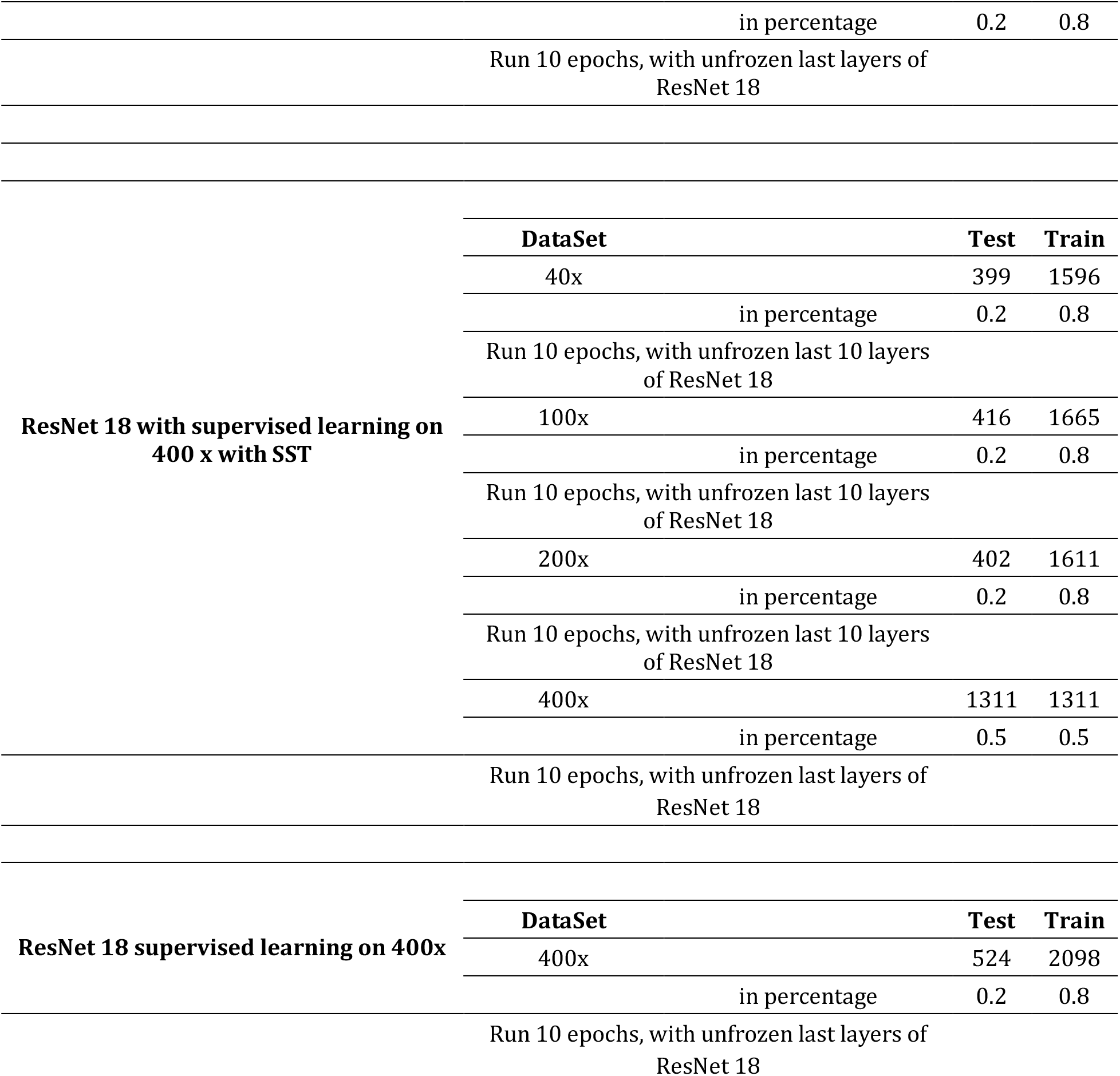
Data set overview

**Table 2.** ResNet 18 with supervised learning on 40 x

|  |  |
| --- | --- |
| Sensitivity | 0.97 |
| Specificity | 0.99 |
| Positive Predictive Value (Precision) | 0.99 |
| Negative Predictive Value | 0.97 |
| False Positive Rate | 0.01 |
| False Discovery Rate | 0.01 |
| False Negative Rate | 0.03 |
| Accuracy | 0.98 |
| F1 Score | 0.98 |
| Matthews Correlation Coefficient | 0.96 |

**Fig2.**
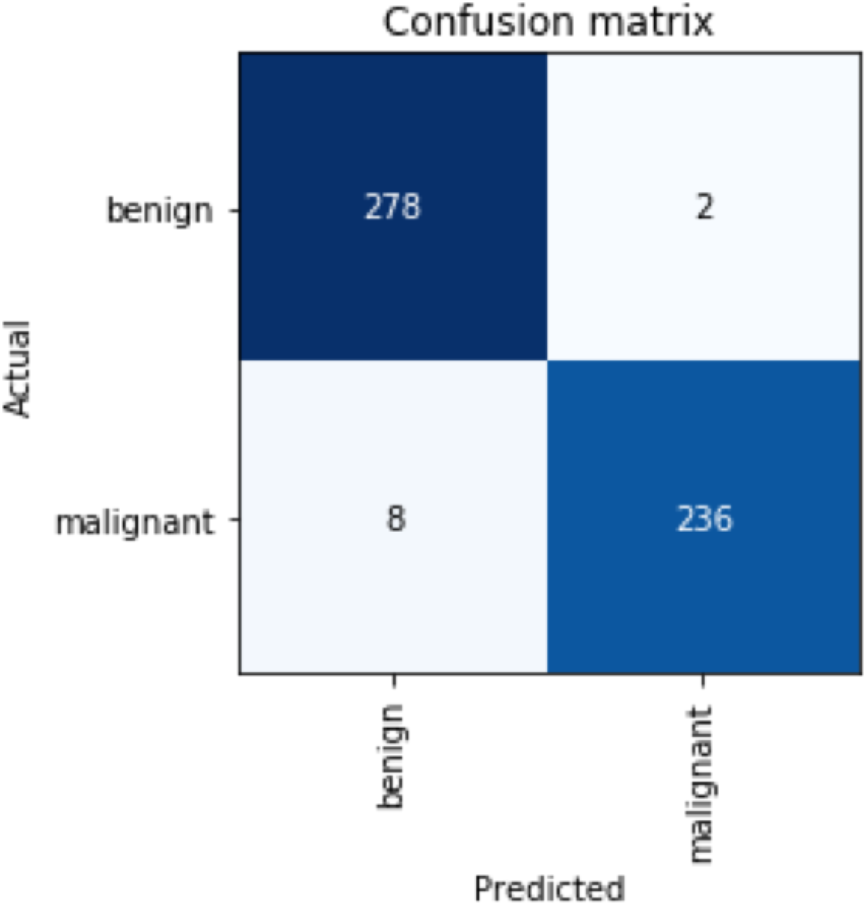
ResNet 18 with supervised learning on 40 x

**Table 3.** ResNet 18 with SST on 400x

|  |  |
| --- | --- |
| Sensitivity | 0.99 |
| Specificity | 1 |
| Positive Predictive Value (Precision) | 1 |
| Negative Predictive Value | 0.99 |
| False Positive Rate | 0 |
| False Discovery Rate | 0 |
| False Negative Rate | 0.01 |
| Accuracy | 1.0 |
| F1 Score | 1.0 |
| Matthews Correlation Coefficient | 0.99 |

**Fig3.**
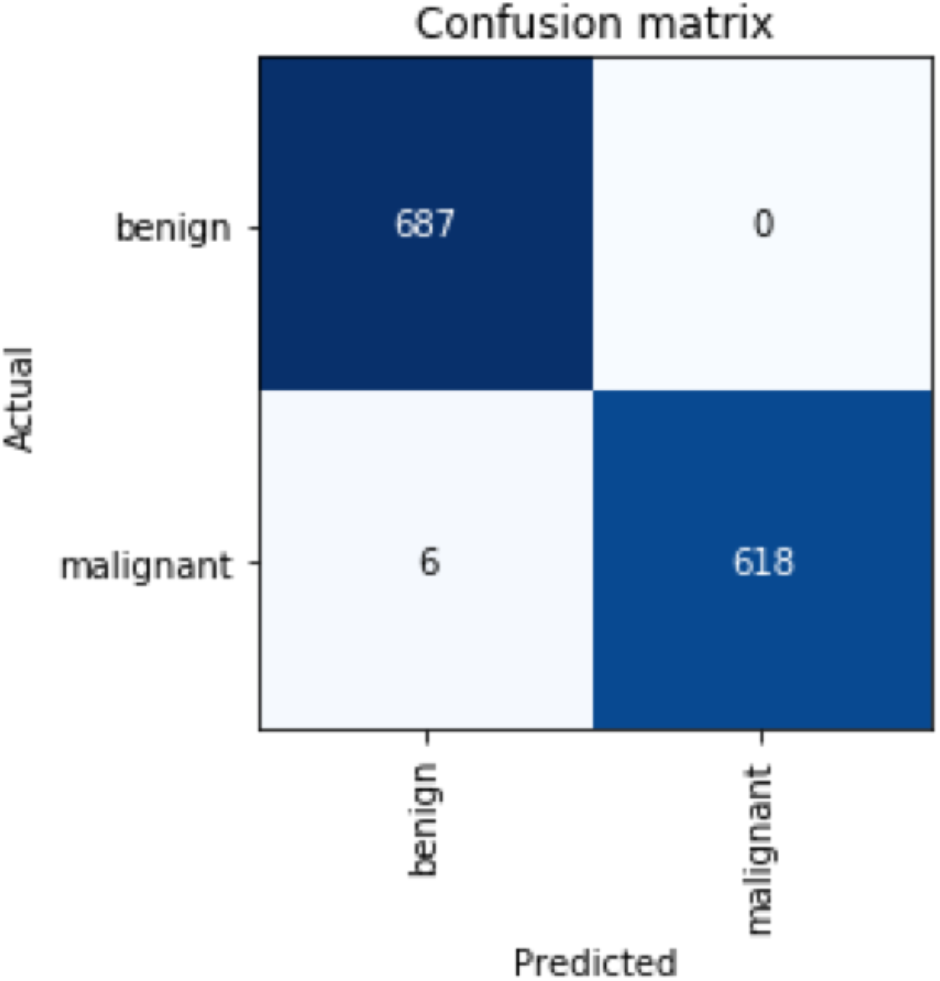
ResNet 18 with SST on 400x

**Fig 4.**
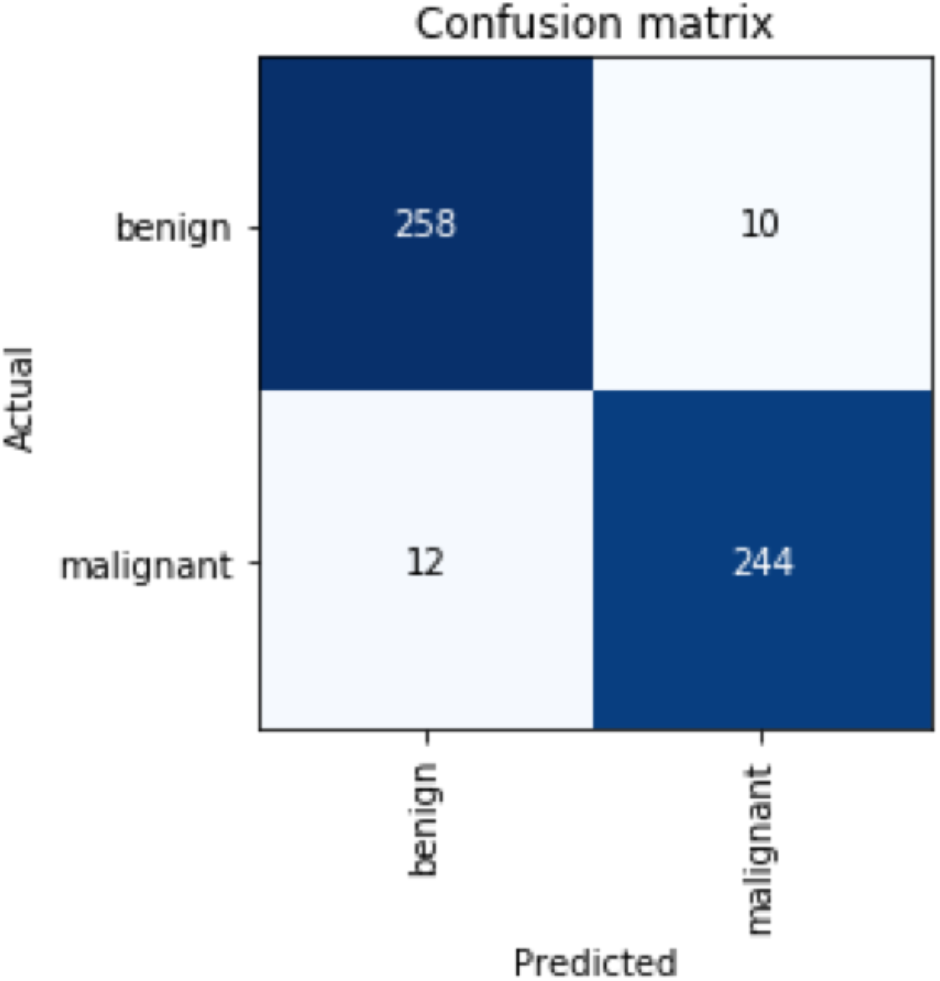
ResNet18 with supervised Learning on 400x

**Table 4.**
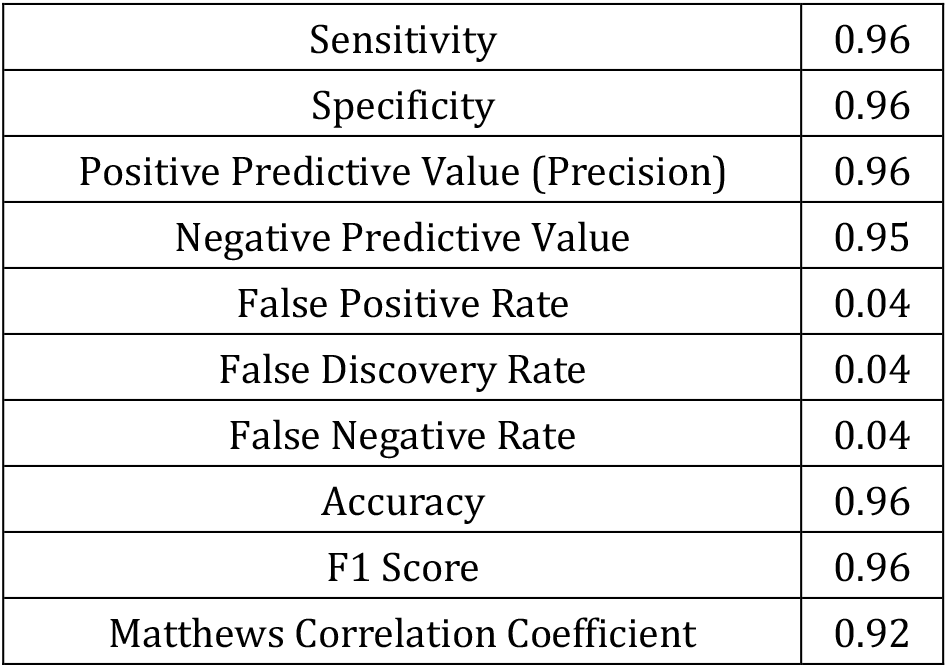
ResNet 18 with supervised learning on 400x

|  |  |
| --- | --- |
| Sensitivity | 0.96 |
| Specificity | 0.96 |
| Positive Predictive Value (Precision) | 0.96 |
| Negative Predictive Value | 0.95 |
| False Positive Rate | 0.04 |
| False Discovery Rate | 0.04 |
| False Negative Rate | 0.04 |
| Accuracy | 0.96 |
| F1 Score | 0.96 |
| Matthews Correlation Coefficient | 0.92 |

## Discussion

In the present study, the analysis of digitized pathology images with the application of convolutional neural network Resnet -18 architecture with SST revealed very encouraging results. A very high F-1 score of 1.0, has been achieved by the ResNet 18 with SST on magnification of 40 as compared to F-1 score of 0.9823, which is achieved by the application of ResNet-18 with supervised learning on 40x. Similarly, a better score (0.9957) of F-1 has been achieved by ResNet-18 with SST on 400x as compared to the F-1 score of 0.9591 by the ResNet 18 with supervised learning on 400x.

The application of SST also solved the problem of images with variable magnifications. The trained network on the magnification of 40x magnification also accurately classified the images of same lesion with magnification other than 40x. The finding would further pave the pathway for application of deep learning for the histological diagnosis of pathological lesions.

The histological examination of tissue is of vital importance for the diagnosis of cancer. The hematoxylin and eosin stained sections are being microscopically examined by the histopathologists for the conclusive diagnosis of malignancies over the past century. With the rising number of cancer patients, it would be very necessary to explore the option of automation in the process of histological examinations as it may increase the speed and accuracy for the diagnosis of cancer.

In the recent past decade, the revolution of digital technology has transformed many fields and it has a healthy impact in the patient care. One of the important aspects of digital technology is the application of artificial intelligence (AI) in health care. AI is used in the speech recognition and for the analysis of the data related to health care.

The machine learning approaches are yielding promising results in the evaluation of radiological images particularly for the diagnosis of malignant tumors [10-12]. Similarly computer vision based systems are being evaluated for the histopathological diagnosis of the surgical biopsy specimens and studies have revealed very good results [13-15].

After the better control of infectious microbial pathogen, the other challenge to the human health is cancer. The clinicians were aware about the clinical behavior of cancer in the ancient times. In 3000 – 2500 BC, the Egyptian physicians described about an incurable disease of breast with the features such as cool to touch, bulging and involvement of whole breast [16]. But the clinicians were unaware of the malignant cell until the compound microscope was developed. Microscopic examination of biologic tissue started in the 17^th^ century but with the development of achromatic lenses, modern era of microscopy began [16].

Friedrich von Esmarch (professor of surgery at Kiel) presented forceful arguments at the German Surgical Congress of 1889 on the need to establish a microscopic diagnosis before operating in suspected cases of malignant tumors requiring extensive mutilating procedures.

At present, the management of cancer includes accurate classification of malignant tumor. Since the breast cancer is the most common cancer among the women all over the world and with the increase in the population size, the number of breast cancer are rising. It would be very important to apply the deep learning technique for the accurate diagnosis and classification of breast cancer as the CNN is being applied for the diagnosis of various lesions on radiological images [17].

## Conclusion

The analysis of digitized pathology images with the application of convolutional neural network Resnet -18 architecture with SST revealed excellent results, which is demonstrated by a very high F-1 score of 1.0.

The development of scale invariant system with application of SST solved the problem of images with variable magnifications. The finding would further pave the pathway for application of deep learning for the histological diagnosis of pathological lesions.

## Data Availability

All data is publicly available

